# Rapid, High-Throughput, Cost Effective Whole Genome Sequencing of SARS-CoV-2 Using a Condensed One Hour Library Preparation of the Illumina DNA Prep Kit

**DOI:** 10.1101/2022.02.07.22269672

**Authors:** Rebecca Hickman, Jason Nguyen, Tracy D. Lee, John R. Tyson, Robert Azana, Frankie Tsang, Linda Hoang, Natalie Prystajecky

## Abstract

The ongoing COVID-19 pandemic necessitates cost-effective, high-throughput, and timely genomic sequencing of SARS-CoV-2 viruses for outbreak investigations, identifying variants of concern (VoC), characterizing vaccine breakthrough infections, and public health surveillance. Additionally, the enormous demand of genomic sequencing on supply chains and the resulting shortages of laboratory supplies necessitate the use of low-reagent and low-consumable methods. Here, we report an optimized library preparation method where the same protocol can be used in a STAT scenario, from sample to sequencer in as little as eight hours, and a high-throughput scenario, where one technologist can perform 576 library preparations over the course of one 8-hour shift. This new method uses Freed et al.’s 1200 bp primer sets (Biol Methods Protoc 5:bpaa014, 2020, https://doi.org/10.1093/biomethods/bpaa014) and a modified and truncated Illumina DNA Prep workflow (Illumina, CA, USA). Compared to the original, application of this new method in hundreds of clinical specimens demonstrated equivalent results to the full-length DNA Prep workflow at 45% the cost, 15% of consumables required (such as pipet tips), 25% of manual hands-on time, and 15% of on-instrument time if performing on a liquid handler, with no compromise in sequence quality. Results suggest that this new method is a rapid, simple, cost-effective, and high-quality SARS-CoV-2 whole genome sequencing protocol.

## INTRODUCTION

Severe acute respiratory syndrome coronavirus 2 (SARS-CoV-2), the causative pathogen for the novel 2019 coronavirus disease (COVID-19), is responsible for the pandemic first identified in Wuhan, China, and since has spread worldwide. As of June 2nd 2021, there have been over 171 million cases and 3.5 million COVID-19 deaths confirmed (https://coronavirus.jhu.edu/), and the pandemic continues to have devastating social, health, and economic impacts globally. The reliance on genomics for the study of viral transmission, identifying variants of interest (VoI) and of concern (VoC), and for studying vaccine breakthrough has necessitated rapid, high-throughput, and high-quality whole genome sequencing. This has resulted in unprecedented demand for laboratory reagents, consumables, equipment, and highly skilled laboratory staff to generate sequence data. With global shortages of laboratory reagents and consumables, there is widespread need for methods with low-reagent and low-consumable requirements that are scalable.

A multiplexed 1200 bp tiled primer amplicon approach previously described (1), which generates high and consistent coverage amplification across the SARS-CoV-2 genome, was chosen for best compatibility with the DNA Prep library preparation kit (Illumina, CA, USA) compared to other widely used primer sets (2, 3). Illumina’s DNA Prep library preparation kit has been shown to generate high-quality, robust library preparations with a method that is scalable and liquid handler compatible. However, this library preparation method is comparatively expensive and more complex than alternatives. Here we use an optimized and truncated Illumina DNA Prep method that minimizes cost, hands-on time, and complexity of work while maintaining high-quality and robust sequence data.

## MATERIALS AND METHODS

### SARS-CoV-2 RNA samples and real-time RT-PCR

A total of 450 clinical samples tested positive for SARS-CoV-2 by various validated laboratory methods throughout the province of British Columbia (BC) were forwarded to the BC Centre for Disease Control Public Health Laboratory (BCCDC PHL) for whole genome sequencing. These samples—collected by either a nasopharyngeal (NP) swab in Yocon transport media or 0.9% saline gargle—were extracted at the BCCDC PHL using Thermofisher’s Viral RNA extraction kit on the MagMAX™/KingFisher™ Flex extractors.

### cDNA synthesis and tiled amplicon generation

cDNA synthesis and amplicon generation were performed as previously described by Freed et al. in steps 1-13 on the protocols.io web-based platform (dx.doi.org/10.17504/protocols.io.bgc8jszw).

### Library preparation and Illumina sequencing

In the originally validated protocol, working in sets of 96 samples, the two multiplex primer pools were combined and purified at a 1:1 volume with AMPure XP beads (Beckman Coulter, IN, USA) and two sequential washes with 80% ethanol. The library preparation was performed with the purified amplicons as per the manufacturer’s recommendations (4), which consists of tagmentation, tagmentation stop, three washes, a reduced 5-cycle library PCR with indexing, and a dual-sided size selection and purification, before individual libraries are pooled (Fig 1A).

**Fig 1.**
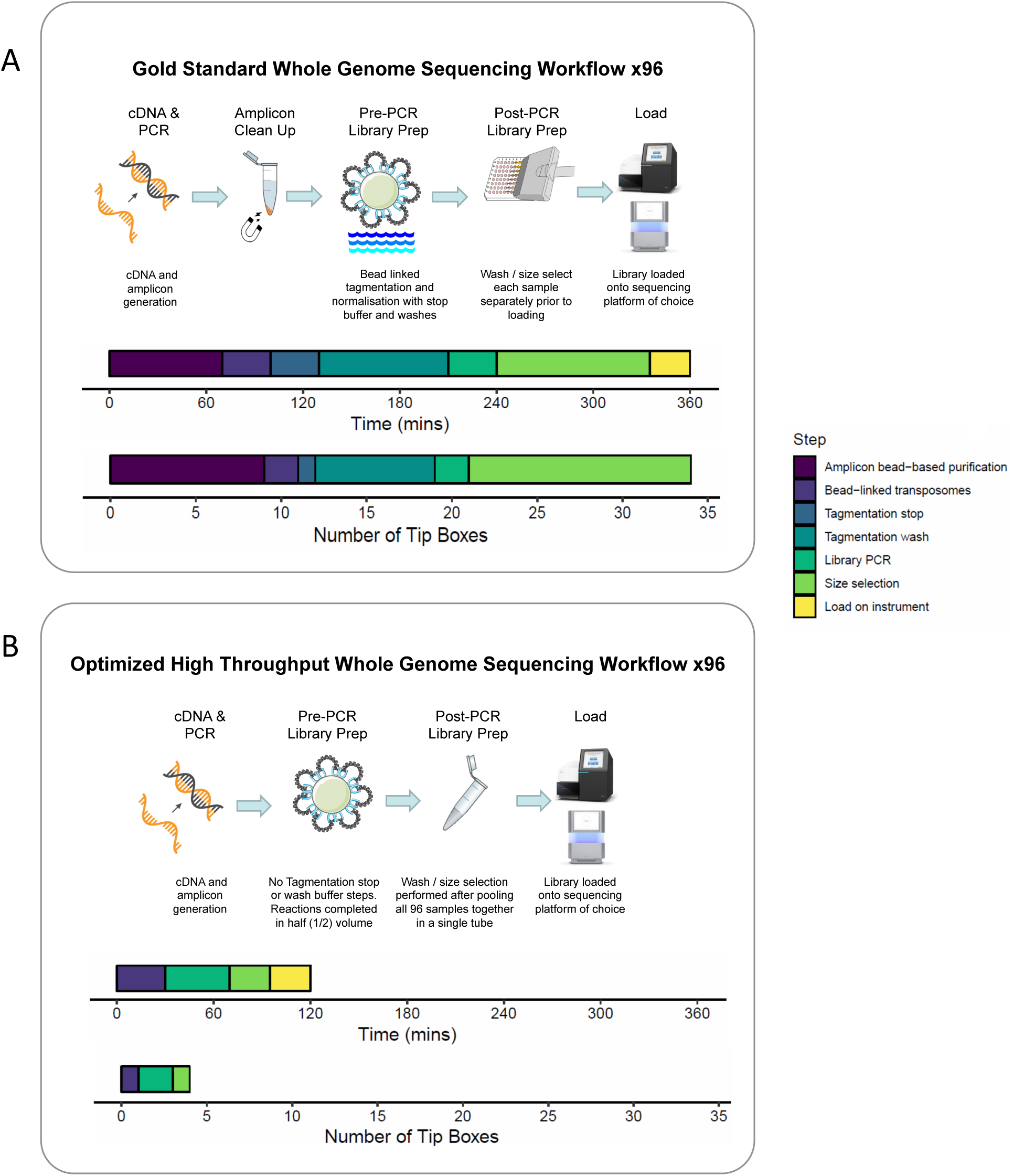
Comparison of step-wise workflow, total time to complete, and tip box usage between the (A) original whole genome sequencing method and the (B) new method (dx.doi.org/10.17504/protocols.io.b3vgqn3w).

The optimized library preparation method, performed at half reaction volumes, eliminates amplicon purification, tagmentation stop, and post-tagmentation washes entirely (Fig 1B). Additionally, pooling was performed on the indexed post-PCR products, rather than the libraries, allowing the dual-sided size selection and purification steps to be performed per 96-sample pool instead of per sample.

Briefly, the multiplex primer pools were combined. Next, 15µl of amplicon, 5µl of bead linked transposomes (BLT) and 5µl of tagmentation buffer 1 (TB1) were heated on a thermal cycler at 55°C for 15 minutes. On a magnet, the supernatant was removed and taken off the magnet, the beads were then re-suspended in 10µl of enhanced PCR mix (EPM), 10µl of ultrapure water, and 5µl of Illumina’s Unique Dual Indexes before being run on a reduced 8-cycle library PCR. After PCR, equal volumes of each library supernatant were pooled. In a single microcentrifuge tube, 45µl of the post-PCR pool was added to 40µl of ultrapure water and 45µl sample purification beads (SPB). After five minutes, 125µl of the supernatant was transferred to a new microcentrifuge tube with 15µl of SPB. After a final 5-minute incubation, two sequential 80% ethanol washes were performed on the magnetic beads before the tagmented, size selected, and purified library was eluted in resuspension buffer (RSB). The finished library was quantified by Qubit, denatured, and diluted to a final concentration of 15pM, and 96 samples were sequenced using the Illumina MiSeq system using a 2 × 150 bp MiSeq Reagent Kit v2 Micro. A detailed laboratory method can be found on protocols.io (dx.doi.org/10.17504/protocols.io.b3vgqn3w).

### epMotion Automation

The five runs for this report were performed manually; however, this protocol has since been automated on the Eppendorf epMotion 5075t liquid handler and has been used to prepare over 50,000 libraries. Using only four boxes of pipet tips, this method is on the instrument for 45 minutes with user intervention only required to transfer the plate to a thermal cycler. The epMotion protocol is available on request.

### Data analysis

The data was analyzed using a modified Nextflow bioinformatics pipeline (https://github.com/BCCDC-PHL/ncov2019-artic-nf) from the Simpson Lab (https://github.com/jts/ncov2019-artic-nf) that is built upon the original Connor laboratory’s ncov2019-artic-nf Nextflow pipeline (https://github.com/BCCDC-PHL/ncov2019-artic-nf). Reports describing sequencing quality metrics—including genome completeness, depth of coverage, and quality flags—and lineage information were generated using ncov-tools from the Simpson Lab (https://github.com/jts/ncov-tools). Lineages were assigned according to the Pangolin version 2.4.2 (5). Resource utilization, estimated in terms of time, cost, and consumables used, was compared between the new and original sequencing methods to determine potential savings.

## RESULTS

### Illumina quality metrics

The basic quality metrics used for Illumina sequencing remained high and met the expected values on sequencing runs using the new method. Over five MiSeq runs, the original method had a mean 94.2% of reads with a Q-score >30 (range 92.7 to 95.0%), compared to the new method which had a mean 95.1% of reads with a Q-score >30 (range 94.2 to 95.8%).

### Sequencing depth of coverage and genome completeness

The mean sequencing depth on the full-length DNA Prep method and the new reduced library preparation method were 412X coverage and 433X coverage, respectively. Using the original method, 387/450 (86.0%) of sequences passed quality metrics, where a pass is >85% of the genome sequenced to >10X depth (Table 1). In comparison, 389/450 (86.4%) sequences passed the quality metrics by the new method. By the original method, 363/450 (80.7%) sequences had over 99% of the SARS-CoV-2 genome and by the new method, 366/450 (81.3%) sequences had over 99% genome coverage (Table 1). Of the 67 samples that did not pass the sequencing metrics on either method, 43 (64.2%) had Ct values available, of which 43/43 (100%) had a Ct value >28 and 41/43 (95.3%) had a Ct value >30. The mean Ct value of a sample that did not pass the quality metrics was 34.3 (Fig 2).

**Table 1.** Genome completeness for each library method

|  | Original method | New method |
| --- | --- | --- |
| Number of samples | 450 | 450 |
| % Passed quality metric (>85%) | 86.0 (387/450) | 86.4 (389/450) |
| % Complete genome (>99%) | 80.7 (363/450) | 81.3 (366/450) |
| % Lineages called (>70%) | 86.8 (391/450) | 87.1 (392/450) |

**Fig 2.**
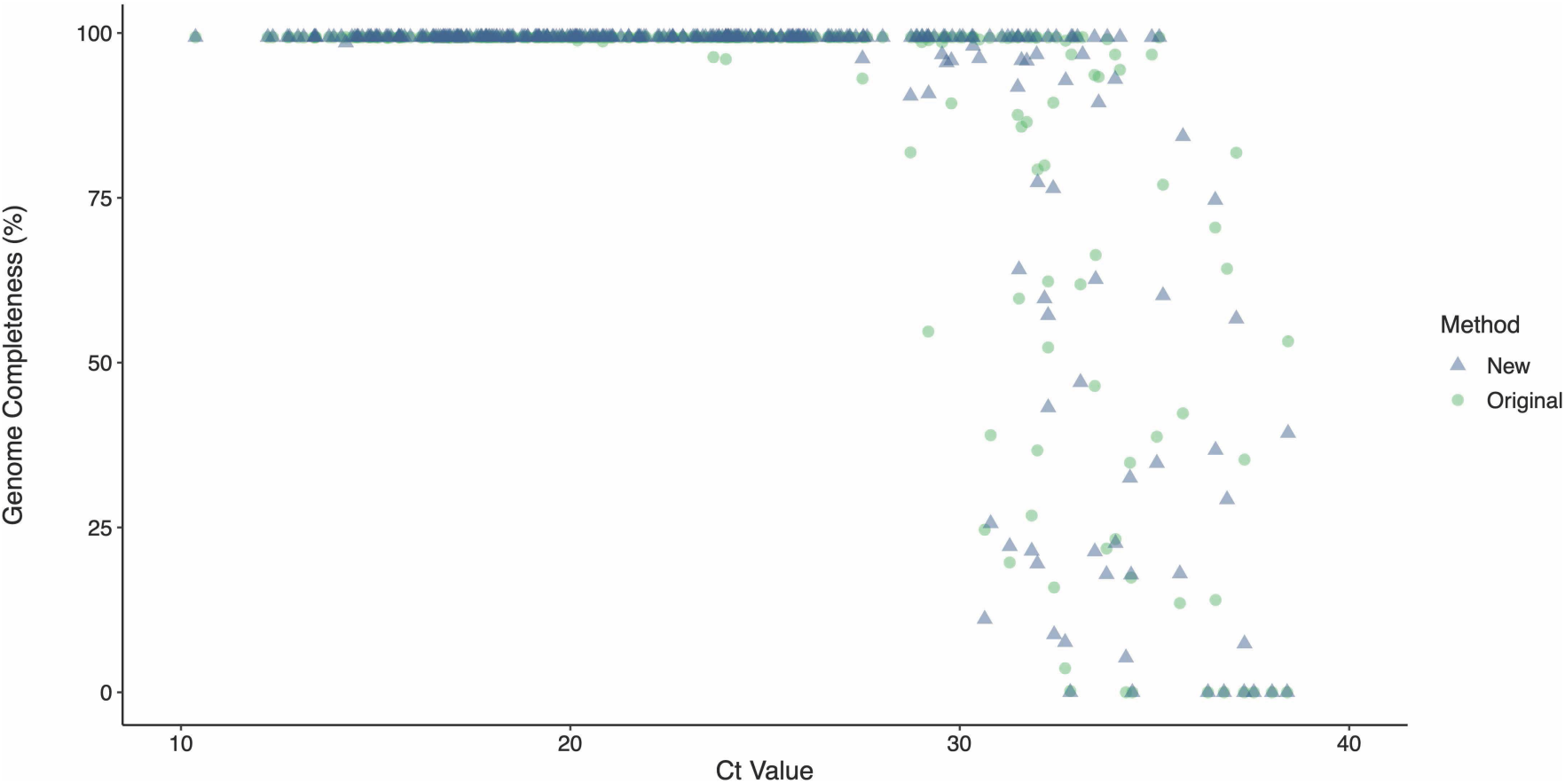
Percent genome completeness for the new method (triangles) and original method (circles) based on available cycle threshold (Ct) values.

### Lineage calls

A >70% genome completeness was required to identify the SARS-CoV-2 lineage. In total, 391/450 (86.8%) of libraries from the original method and 392/450 (87.1%) of libraries from the new method had sufficient genome completeness to assign a lineage (Table 1). In the 386 samples that had a lineage assignment by both methods as well as a passing >85% coverage and no quality flags, 383 (99.2%) of lineages matched between methods. The three samples with lineage mismatches had 3-13% of their genomes unsequenced, with the missing locations being key mutations sites that altered the lineage calls.

### Resource utilization

The new method required less total time to complete compared to the original method, both when performed manually (2 hours vs. 6 hours, respectively) and on an Eppendorf epMotion 5075t instrument (1.5 hours vs. 6.5 hours, respectively) (Table 2, Fig 1B). The new method was also associated with a lower cost of library preparation ($31 vs. $68) and required fewer tip boxes (4 vs. 34) compared to the original method (Table 2, Fig 1B).

**Table 2.** Time, cost, and consumable savings

|  | Original method | New method |
| --- | --- | --- |
| Hands-on time (manual) | 4 hours, 30 minutes | 1 hour |
| Total time to completion (manual) | 6 hours | 2 hours |
| Time on instrument (epMotion) | 6 hours | 45 minutes |
| Total time to completion (epMotion) | 6 hours, 30 minutes | 1 hour, 30 minutes |
| Cost of library prep (CAD list price) | \$68 | \$31 |
| Number of tip boxes | 34 | 4 |

## DISCUSSION

Sequencing has played an invaluable role in the response to the COVID-19 pandemic. Specifically, sequencing has been used to complement outbreak investigations, to study viral transmission and introductions, to identify emerging variants, and to understand vaccine breakthrough infection. Ongoing work in this area, however, demands optimization of laboratory work for whole genome sequencing in order to increase sequencing capacity, improve turn-around-time, and reduce cost without compromising sequence quality. This report describes a new sequencing method that incorporates workflow modifications that save time, limit the use of reagents and consumables, and maintain high-quality sequencing results.

### Time savings

The original Illumina DNA Prep library preparation, performed manually, requires almost a full 8-hour work shift to complete, the vast majority of it being hands-on time. In addition, the multiple wash steps are difficult on the laboratory worker and prone to error. The original method was programmed on an Eppendorf epMotion 5075t, but took 6.5 hours on the instrument to complete. Both of these options were slow and laborious for the laboratory workers and instruments. The new method removes amplicon purification, tagmentation washes, and pools the dual sided size selection into a single tube thereby removing the vast majority of the pipetting steps. A single laboratory worker is now capable of producing 576 library preparations by this method manually in an 8-hour shift. Furthermore, when performed on the epMotion instrument, the entire method takes only 45 minutes for a 96-well plate, freeing the instrument for the next library preparation and allowing one instrument to perform 7+ 96-well plate libraries in an 8-hour shift. With the same instruments and staff, we were able to sequence 6-8X as many libraries as the previous method in the same amount of time. This is particularly important during staff shortages or during testing surges when human resources are a limiting factor.

In addition, as the library preparation takes one hour, it allows for the unique opportunity of going from sample to sequencer in the course of one shift in a STAT scenario. Extraction, cDNA synthesis, and amplicon generation typically take around six hours; however, our reduced library preparation method allows the samples to be on the sequencer by the end of the shift.

### Reagent and consumable savings

During a time of global laboratory reagent and consumable shortages, the elimination of multiple steps from the original protocol has other positive effects. Removing the amplicon clean up and halving the reaction volume of the DNA Prep library preparation reduced the reagent cost of whole genome sequencing by nearly 55%, making an initially expensive library preparation kit more affordable. Additionally, removing the majority of steps, especially washes, reduces pipet tip usage substantially from 34 tip boxes to four (an 85% reduction) for a 96-sample library preparation.

### High quality results

Despite the removal of the amplicon purification and the majority of library preparation steps, results have shown that the quality of the sequencing was not compromised. The percent of samples with complete genomes (>99%) and passing quality control cutoffs (>85% genome completeness) were equivalent between the two methods. Of samples passing quality control, 99.2% of the 450 samples had a matching lineage between the methods; the three that did not have matching lineages had 3-13% of the genome missing in key lineage-defining sites.

In conclusion, this study demonstrates a rapid library preparation method for SARS-CoV-2 whole genome sequencing that produces consistent and high-quality data with equivalent results to the original full length DNA Prep library preparation as described from the manufacturer. With an overall savings of >75% hands-on time at 45% the cost and using 15% of the consumables compared to a typical DNA Prep library preparation, this new protocol is an efficient, scalable, and pragmatic alternative for SARS-CoV-2 whole genome sequencing.

## Data Availability

All data produced in the present study are available upon reasonable request and with permission of the data steward.

## AUTHORS’ CONTRIBUTIONS

RH designed the experiment, tested the protocol, analyzed the data, and wrote the manuscript. JN conceived the idea for the high throughput method, assisted with the development, testing, and automation of the protocol, and helped with analyzing the initial developmental data. TDL and JT contributed to experimental design. FT and RA provided operational support and coordination for implementation of the protocol. NP and LH provided study oversight. NP, JT, and TDL critically reviewed the manuscript and all authors approved the final draft.

## ACKNOWLEDGEMENTS

We would like to thank the Virology Laboratory at the BCCDC PHL for performing routine diagnostic testing of SARS-CoV-2. We would like to thank the Bacteriology and Mycology Laboratory at the BCCDC PHL for performing the routine whole genome sequencing of SARS-CoV-2. We would also like to the BCCDC data analytics and bioinformatics team for pipeline design and optimization, and Jessica Caleta for assistance with figure design.

## ETHICS STATEMENT

This study was conducted under the auspices of a quality improvement initiative, authorized by the Provincial Health Officer under the Public Health Act. Ethical oversight was waived by the University of British Columbia Clinical Research Ethics Board, in accordance with Article 2.5 of the Tri-Council Policy Statement 2 (TCPS 2).

